# Elevated blood pressure in the emergency department – a risk factor for incident cardiovascular disease: An EHR-based cohort study

**DOI:** 10.1101/19004382

**Authors:** P. Oras, H. Häbel, P. H. Skoglund, P. Svensson

## Abstract

**Objectives:** In the emergency department (ED), high blood pressure (BP) is commonly observed but mostly used to evaluate patients’ health in the short-term. We aimed to study whether ED-measured BP is associated with incident atherosclerotic cardiovascular disease (ASCVD), myocardial infarction (MI), or stroke in long-term, and to estimate the number needed to screen (NNS) to prevent ASCVD.

**Design:** Electronic Health Records (EHR) and national register-based cohort study. The association between BP and incident ASCVD was studied with Cox-regression.

**Setting:** Two university hospital emergency departments in Sweden.

**Data sources:** BP data were obtained from EDs EHR, and outcome information was acquired through the Swedish National Patient Register for all participants.

**Participants:** All patients ≥18 years old who visited the EDs between 2010 to 2016, with an obtained BP (n=300,193).

**Main outcome measures:** Incident ASCVD, MI, and stroke during follow-up.

**Results:** The subjects were followed for a median of 42 months. 8,999 incident ASCVD events occurred (MI: 4,847, stroke: 6,661). Both diastolic and systolic BP (SBP) was associated with incident ASCVD, MI, and stroke with a progressively increased risk for SBP within hypertension grade 1 (HR 1.15, 95% CI 1.06 to 1.24), 2 (HR 1.35, 95% CI 1.25 to 1.47), and 3 (HR 1.63, 95% CI 1.49 to 1.77). The six-year cumulative incidence of ASCVD was 12% for patients with SBP ≥180 mmHg compared to 2% for normal levels. To prevent one ASCVD event during the median follow-up, NNS was estimated to 151, whereas NNT to 71.

**Conclusions:** BP in the ED is associated with incident ASCVD, MI, and stroke. High BP recordings in EDs should not be disregarded as isolated events, but an opportunity to detect and improve treatment of hypertension. ED-measured BP provides an important and under-used tool with great potential to reduce morbidity and mortality associated with hypertension.

## INTRODUCTION

High blood pressure (BP) is a well-studied and established risk factor for cardiovascular disease (CVD)^1-3^ and is commonly observed in the emergency department (ED).^4^ Hypertension accounts for nearly fifty per cent of the mortality in stroke and heart disease, and every year approximately 9.4 million cardiovascular deaths occur due to complications of hypertension, which makes hypertension the most important risk factor for disability and death worldwide.^5^

Finding and treating patients with hypertension is a priority in CVD prevention. Studies have shown that for every 10-20-mmHg reduction in systolic blood pressure (SBP), there is a substantial risk reduction for CVD,^6-8^ and that patients with higher BP levels benefit from more intensive drug intervention.^9^ However, a majority of the patients with high BP do not achieve BP control (BP < 140/90 mm Hg), and many are unaware of their condition.^10^ In the ED, BP is measured on almost every patient in order to assess the patient’s condition and acuity^11,12^ regardless of the presenting chief complaint. Previous research has reported that up to one-third of the patients in the ED have BP levels above the threshold for hypertension, and severely elevated levels have been observed in 20 % of the cases.^11^ Elevated BP in the ED can be explained by factors such as pain, anxiety or other stressors, which raises the question of its predictive value and if hypertension screening based on routine recordings in the ED are useful.^4^

Although the value of BP in the ED as hypertension screening can be questioned,^13^ it is not known if high BP in the ED is associated with incident atherosclerotic cardiovascular disease (ASCVD), nor the benefit on acting in this setting. Therefore, we aimed to examine if the first measured BP in the ED is associated with incident ASCVD, MI, or stroke during a long-time follow-up. We also aimed to assess the potential benefits of screening for patients with hypertension in the ED by estimating the number needed to screen and treat to prevent ASCVD.

## MATERIALS AND METHODS

### Study design and setting

This electronic health records (EHR) and register-based cohort study included all patients who visited the EDs at Karolinska University Hospital in Solna and Huddinge, Sweden during the period from January 2010 until March 2016, with a BP measured in the ED. The exposure BP was obtained from EDs EHR, and all outcome information were acquired by linkage to national registers for all included participants. The participants were followed regarding incident disease, death or censoring until end of the study on 31st December 2016. The study was registered at ClinicalTrials.gov: NCT03954119.

### Inclusion criteria

The study cohort was all patients visiting the EDs with a BP obtained at arrival. Patients were eligible for inclusion in the study disregarded of the chief complaint as long as they had a BP recorded and were older than 18 years old. Patients without a Swedish personal identification number were excluded since no outcome information could be obtained from these patients. Only the first visit in the ED with a measured BP was included. Recurrent BP measurements during hospitalisation or recurrent visits to the ED with new BP recordings were disregarded.

### Data sources

BP data and all covariates were acquired through Karolinska University Hospital’s EHR for each patient’s ED visit. The National Board of Health and Welfare (NBHW) provided data for outcome events and event dates until the end of December 2016, from the National Patient Register for the selected subjects. Hypertension diagnosis before index visits were collected from this register from the period January 1997 to December 2016. The cause of death and date of death were obtained through the NBHW’s Cause of Death Register. Data for each subject in the registers were merged to one dataset by the unique pseudonymised identification number. Data for pick-up of prescribed drugs were acquired from the NBHW’s Prescribed Drugs Register.

### Exposure definitions

The exposures were defined as the first measured SBP and DBP in the ED; both measured as a part of the triage system according to clinical routine.^12^ BP was categorised into groups due to no certain linear association for BP with the outcomes. The categories were based on the *ESC and ESH* classification of BP grades and the definition of hypertension grades.^14^ SBP categories were defined as <90 mmHg, 90-119 mmHg, 120-129 mmHg, 130-139 mmHg, 140-159 mmHg, 160-179 mmHg, and >179 mmHg. DBP were categorised into <60 mmHg, 60-79 mmHg, 80-84 mmHg, 85-89 mmHg, 90-99 mmHg, 100-109 mmHg, and >110 mmHg (Supplementary Table 1).

### Endpoint definitions

The composited primary endpoint was ASCVD, defined as the first occurring event of CHD death (fatal MI and sudden cardiac death), non-fatal MI and all-cause stroke, based on the pooled cohort risk equations.^15^ Secondary endpoints were fatal and non-fatal MI, all-cause stroke, ischemic stroke, and intracerebral haemorrhage. Data for endpoints were derived as the primary diagnosis from the National Patient Register and Cause of Death Register, according to the *International Classification of Diseases 10*^*th*^ *edition*^16^ (ICD-10). ICD-10 classification for ASCVD was fatal CHD (MI: I210-I214, I219, I220, I221, I228, I229, sudden cardiac death: I461 or I469), non-fatal MI (I210-I214, I219, I220, I221, I228 or I229), fatal and non-fatal ischemic stroke (I630-I635, I638 or I639) (Supplementary Table 2). Only the first event that occurred in the follow-up period was included in the endpoint ASCVD, and all following recurrent events were disregarded. Secondary endpoints were defined as fatal and non-fatal MI (I210-I214, I219, I220, I221, I228 or I229. I200 were included if combined with intervention procedure codes FNA-FNG, FNW), all-cause stroke (I610-I619, I630-I635, I638-I639, or I649), ischemic stroke (I630-I635 or I638-I639), and intracerebral haemorrhage (I610-I619) (Supplementary Table 3).

### Variables for subgroup analyses

Patients with a diagnosis of hypertension before the ED visit were identified through a record of hypertension (I109) or a pick-up of a prescription of antihypertensive drugs within the preceding 12 months of the index visit. Antihypertensive drugs were defined according to the anatomic therapeutic classification (ATC) (Supplementary Table 4).

### Follow-up period

The baseline for follow-up was defined as the time patients were discharged either from the ED or from inpatient care, in order to not include events during hospital care linked to the index visit and thereby avoid effects of an outcome occurring before the exposure. For patients that were not admitted to inpatient care but were directly discharged from the ED, one day was added after they were discharged, in order to avoid misclassification of a hospitalisation that had been referred to from the ED-visit.

### Addressing potential bias

The risk of selection bias was minimised by the broad inclusion criteria for this study. BP categories were defined according to the clinically relevant definitions by the *ESC and ESH*.^14^ All outcomes were determined in advance with established definitions, disclosed in the trial registration at *clinicaltrials*.*gov*. The Swedish National Patient Register, from which the outcome data were obtained, has high validity.^17^

### Statistical analysis

Hazard ratios (HR) for primary and secondary endpoints were estimated with Cox proportional hazard regression. Age, sex, inclusion year, and hospital were adjusted for in multivariate analyses. Age was controlled with restricted cubic splines with four knots in order to adjust for the gradually increasing impact of higher age on event rates. The reference BP categories in the analysis were set to the normal BP level according to *ESC and ESH* classification;^14^ SBP reference category was 120-129 mmHg, and DBP reference category was 80-84 mmHg. The proportional hazards assumption was evaluated by analysing Schoenfeld residuals. Subgroup analyses for the outcome ASCVD were performed with Cox proportional hazard regression for patients with a history of hypertension, for hospitalised patients, and by chief complaints. All chief complaints were categorised by medicine discipline based on the managing emergency department and type of chief complaint (Supplementary Table 5). Cumulative incidence functions for SBP categories were estimated for the endpoints with the Kaplan-Meier estimator. Cases with missing exposure and outcome data were excluded from the analyses. Descriptive statistics of the study population were presented as mean ± standard deviation (SD) or relative and absolute frequencies grouped by SBP categories. A *p*-value of <0,05 was considered to be significant. STATA version 15.1 was used for the analysis. Participants with missing BP data or had an outcome before follow-up baseline were disregarded from the analysis.

To assess the potential benefit of screening for high BP among patients in the ED, a number needed to screen (NNS) and number needed to treat (NNT) were estimated for CHD, stroke, and ASCVD. The estimates were based on the relative risk reduction (RRR) reported in previous research, where a 10-mmHg SBP reduction yielded a risk reduction by 22% for CHD and 14% for stroke, irrespectively of baseline BP.^8^ Potentially preventable events (PPE) per ED patient were calculated for all subjects with a SBP ≥140 mmHg based on the RRR. PPE was weighted by the visits for each year, and NNS was derived by the inverse of the PPE per ED patient, presented with a 95% CI.

### Patient and public involvement

Participants were not involved in the design of this study, nor the definition of exposures, outcome measures or the setting. Participants were not asked to advise on interpreting the data or contributed to writing the manuscript.

## RESULTS

A total of 300,193 patients were included in the study after the inclusion criteria (Figure 1). The mean age was 50.0±20.2 years, female patients comprised 53.9% (n=161,707) of the study population, and the mean SBP was 139.9 mmHg (Table 1). Subjects were followed for a mean time of 3.5 years, which resulted in 1.1 million person-years. The most common chief complaint was abdominal pain, chest pain, breathing difficulties, headache, and fever. (Supplementary Table 5). SBP data were missing for 59 (0.02%) subjects, and 1,992 (0.67%) subjects had data missing for DBP. No subject were missing data for both SBP and DBP. Outcome data were obtained for all included participants.

**Table 1.** General characteristics of the study population.

|  | Systolic Blood Pressure (mmHg) |  |  |  |  |  |  |  |
| --- | --- | --- | --- | --- | --- | --- | --- | --- |
|  | Total<br>n=300,193 | <90<br>n=1,524 | 90-119<br>n=52,493 | 120-129<br>n=51,667 | 130-139<br>n=55,012 | 140-159<br>n=80,770 | 160-179<br>n=37,207 | ≥180<br>n=21,379 |
| **Age** |  |  |  |  |  |  |  |  |
| Age, mean (SD), y | 50.0 (20.2) | 60.4 (21.6) | 43.0 (19.3) | 43.0 (18.5) | 45.7 (18.9) | 52.0 (19.5) | 60.7 (17.8) | 67.7 (14.8) |
| <65 No. (%) | 219,189 (73.0) | 748 (49.1) | 43,684 (83.2) | 43,790 (84.7) | 44,685 (81.2) | 57,354 (71.0) | 20,433 (54.9) | 8,391 (39.2) |
| 65-74 No. (%) | 38,121 (12.7) | 314 (20.6) | 3,729 (7.1) | 3,689 (7.1) | 5,020 (9.1) | 11,487 (14.2) | 8,121 (21.8) | 5,757 (26.9) |
| >74 No. (%) | 42,883 (14.3) | 462 (30.3) | 5,080 (9.7) | 4,198 (8.1) | 5,307 (9.6) | 11,929 (14.8) | 8,653 (23.3) | 7,232 (33.8) |
| **Sex** |  |  |  |  |  |  |  |  |
| Female, No. (%) | 161,707 (53.9) | 864 (56.7) | 36,588 (69.7) | 30,353 (58.7) | 27,515 (50.0) | 36,768 (45.5) | 17,546 (47.2) | 12,002 (56.1) |
| **Blood Pressure** |  |  |  |  |  |  |  |  |
| SBP mean (SD), mmHg | 139.9 (24.2) | 80.4 (8.0) | 110.1 (6.8) | 123.9 (3.1) | 133.9 (3.1) | 147.6 (5.9) | 167.4 (5.9) | 194.6 (14.6) |
| DBP mean (SD), mmHg | 80.8 (15.0) | 54.9 (14.2) | 68.5 (11.3) | 75.0 (10.2) | 79.2 (11.6) | 84.6 (12.1) | 91.0 (14.5) | 98.7 (14.4) |
| **Medical history** |  |  |  |  |  |  |  |  |
| Hypertension, No. (%) | 75,385 (25.1) | 663 (43.5) | 7,977 (15.2) | 7,410 (14.3) | 9,728 (17.7) | 21,915 (27.1) | 15,551 (41.8) | 12,111 (56.6) |
| Diabetes, No. (%) | 20,526 (6.8) | 152 (10.0) | 2,353 (4.5) | 2,282 (4.4) | 3,017 (5.5) | 6,075 (7.5) | 3,962 (10.6) | 2,675 (12.5) |
| CVD, No. (%) | 13,732 (4.6) | 164 (10.8) | 1,941 (3.7) | 1,638 (3.2) | 1,959 (3.6) | 3,893 (4.8) | 2,388 (6.4) | 1,747 (8.2) |
| **Admission to inpatient care** |  |  |  |  |  |  |  |  |
| Admitted, No. (%) | 76,390 (25.5) | 989 (64.9) | 13,836 (26.4) | 11,293 (21.9) | 12,071 (21.9) | 19,861 (24.6) | 10,858 (29.2) | 7,460 (34.9) |
| **Chief complaints by medical discipline** |  |  |  |  |  |  |  |  |
| Internal Medicine, No (%) | 112,264 (37.4) | 772 (50.7) | 19,246 (36.7) | 18,400 (35.6) | 20,160 (36.6) | 30,328 (37.5) | 14,716 (39.6) | 8,595 (40.2) |
| Surgery, No (%) | 83,956 (28.0) | 350 (23.0) | 17,523 (33.4) | 16,133 (31.2) | 16,038 (29.2) | 21,501 (26.6) | 8,440 (22.7) | 3,935 (18.4) |
| Otolaryngology, No (%) | 13,622 (4.5) | 46 (3.0) | 1,832 (3.5) | 2,163 (4.2) | 2,688 (4.9) | 3,580 (4.4) | 1,938 (5.2) | 1,379 (6.4) |
| Neurology, No (%) | 37,071 (12.3) | 79 (5.2) | 4,785 (9.1) | 5,729 (11.1) | 6,401 (11.6) | 10,626 (13.2) | 5,486 (14.7) | 3,956 (18.5) |
| Orthopaedics, No (%) | 30,819 (10.3) | 89 (5.8) | 4,403 (8.4) | 5,220 (10.1) | 5,730 (10.4) | 9,294 (11.5) | 4,068 (10.9) | 1,993 (9.3) |
| Unknown, No (%) | 22,461 (7.5) | 188 (12.3) | 4,704 (9.0) | 4,032 (7.8) | 3,995 (7.3) | 5,441 (6.7) | 2,559 (6.9) | 1,530 (7.2) |

**Abbreviations** SBP = systolic blood Pressure. DBP = diastolic blood pressure. SD = standard deviation. No = number. y = years.

**Figure 1.**
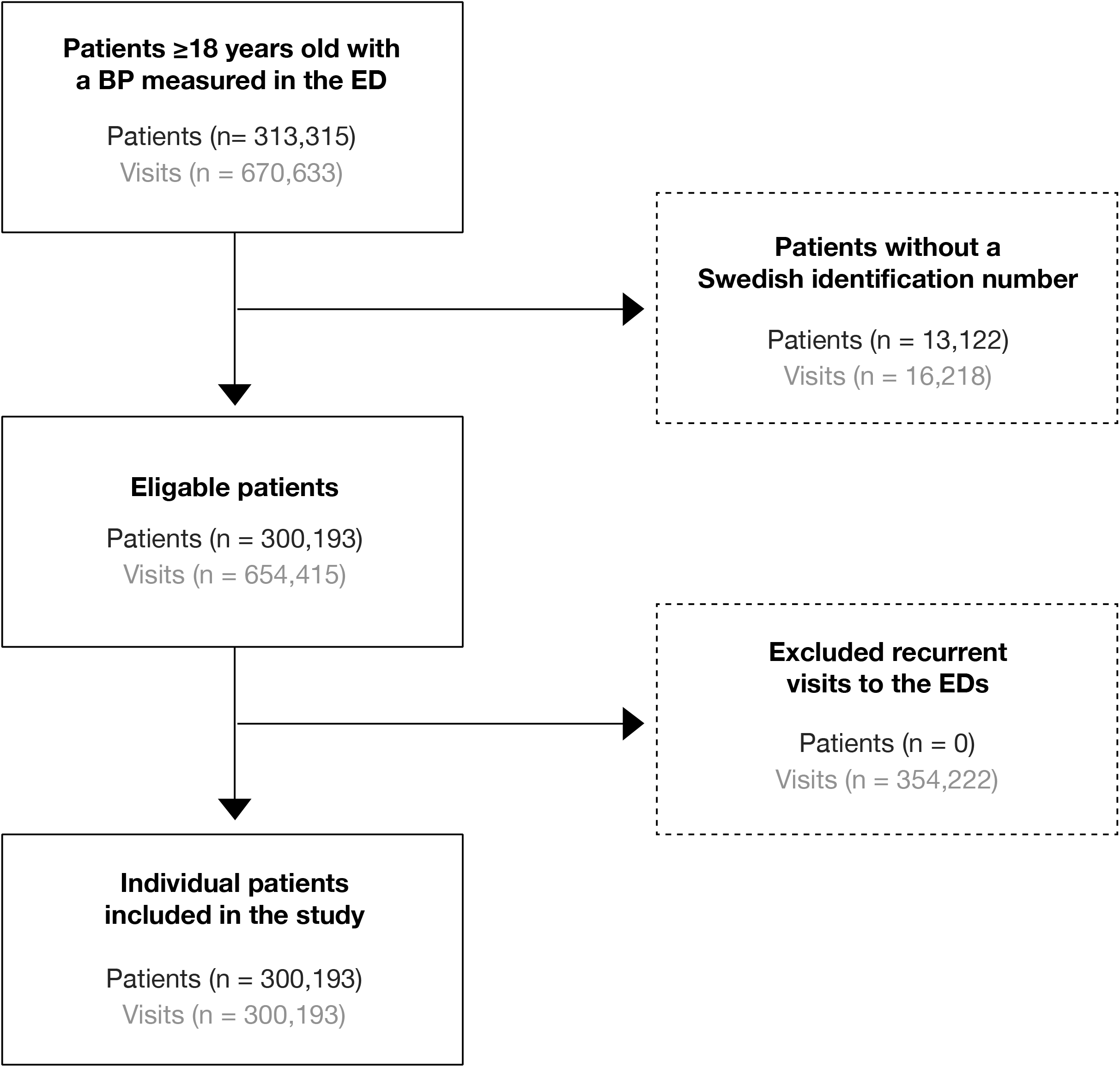
Flowchart of patient selection.

Incident ASCVD occurred in 3.0% (n=8,999) of the cohort during a median follow-up for 42 months (range 0-84 month). Men were more likely to develop ASCVD (3.7%) compared to females (2.4%). BP was associated with incident ASCVD with a progressively increased hazard ratio among patients with SBP above reference categories. Within SBP, the strongest association was observed for ≥180 mmHg (HR 1.63, 95% CI 1.49-1.77) (Table 2). The six-year cumulative incidence of ASCVD for the reference SBP group (120-129 mmHg) was 2% compared to 12% in the highest SBP category (≥180 mmHg) (Figure 2). A statistically significant increased association were observed in all categories above the reference category for DBP.

**Table 2.** Hazard ratio for ASCVD.

| SBP (mmHg) | Crude |  |  | Adjusted <sup>a</sup> |  |  |
| --- | --- | --- | --- | --- | --- | --- |
|  | HR | 95% CI | p-value | HR | 95% CI | p-value |
| <90 | 2.17 | 1.59 to 2.97 | <0.0001 | 0.92 | 0.67 to 1.26 | 0.618 |
| 90-119 | 1.07 | 0.97 to 1.17 | 0.191 | 1.10 | 1.00- to .22 | 0.043 |
| 120-129 <sup>b</sup> | Reference |  |  | Reference |  |  |
| 130-139 | 1.30 | 1.19 to 1.42 | <0.0001 | 1.05 | 0.96 to 1.15 | 0.257 |
| 140-159 | 2.08 | 1.92 to 2.25 | <0.0001 | 1.15 | 1.06 to 1.24 | 0.001 |
| 160-179 | 3.57 | 3.29 to 3.88 | <0.0001 | 1.35 | 1.25 to 1.47 | <0.0001 |
| $\geq 180$ | 5.45 | 5.01 to 5.93 | <0.0001 | 1.63 | 1.49 to 1.77 | <0.0001 |
| DBP (mmHg) | Crude |  |  | Adjusted <sup>a</sup> |  |  |
|  | HR | 95% CI | p-value | HR | 95% CI | p-value |
| <60 | 1.20 | 1.07 to 1.36 | 0.002 | 1.08 | 0.96 to 1.22 | 0.199 |
| 60-79 | 0.89 | 0.83 to 0.95 | <0.0001 | 1.01 | 0.95 to 1.08 | 0.704 |
| 80-84 <sup>b</sup> | Reference |  |  | Reference |  |  |
| 85-89 | 1.20 | 1.11 to 1.30 | <0.0001 | 1.13 | 1.05 to 1.23 | 0.002 |
| 90-99 | 1.43 | 1.33 to 1.53 | <0.0001 | 1.15 | 1.07 to 1.23 | <0.0001 |
| 100-109 | 1.94 | 1.79 to 2.11 | <0.0001 | 1.40 | 1.29 to 1.52 | <0.0001 |
| $\geq 110$ | 2.61 | 2.36 to 2.88 | <0.0001 | 1.71 | 1.55 to 1.89 | <0.0001 |

<sup>a</sup> Adjusted for age, sex, inclusion year, hospital.
<sup>b</sup> Reference category for Cox proportional hazard analysis
**Abbreviations** ASCVD = atherosclerotic cardiovascular disease. SBP = systolic blood pressure. DBP = diastolic blood pressure. HR = hazard ratio. CI = confidence interval.

**Figure 2.**
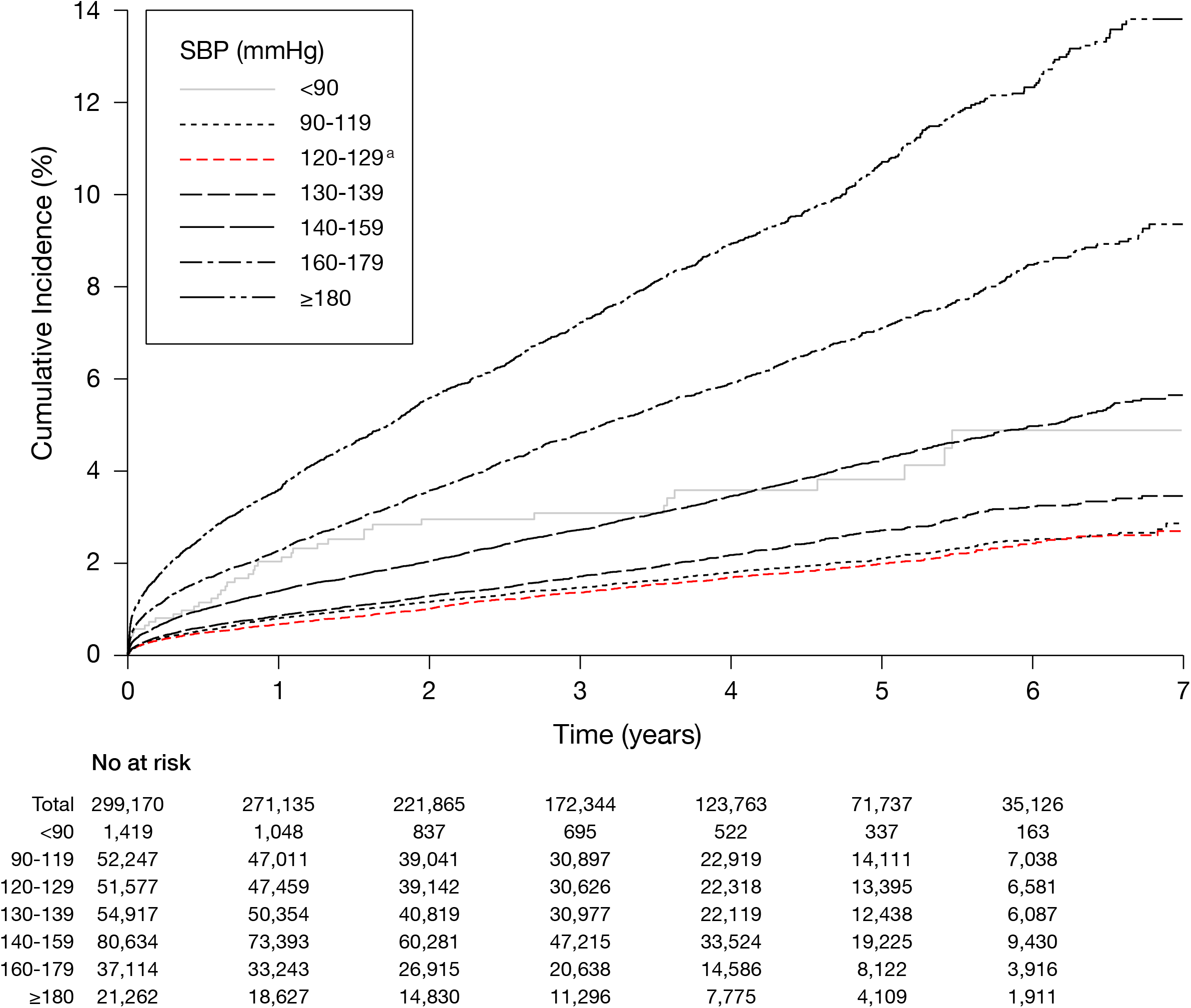
Cumulative incidence of ASCVD. Cumulative incidence are presented by SBP categories. Number at risk presents the number of subjects entering each interval. ^a^ Reference category for Cox proportional hazard regression **Abbreviations** ASCVD: atherosclerotic cardiovascular disease. SBP: systolic blood pressure

For the secondary endpoint MI, a total of 4,847 (1.6%) events occurred in the entire cohort, the majority 63.2% (n=3,117) among men. There was a statistically significant association between BP and incident MI with a gradually increased HR by higher BP above the reference category. (Figure 3-4). The six-year cumulative incidence was 1% for the reference category compared to 7% in the highest SBP group (Figure 5).

**Figure 3.**
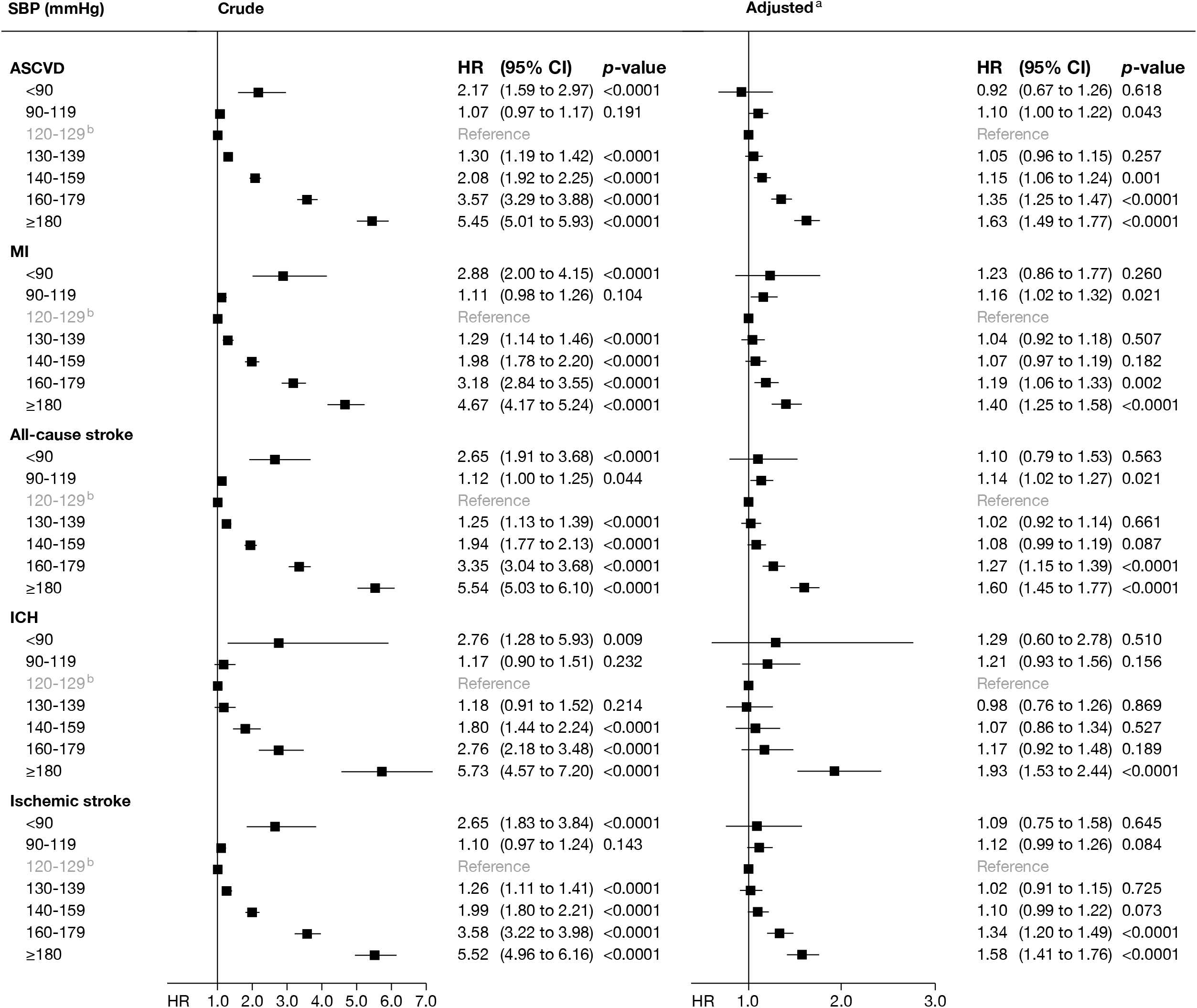
Hazard ratio for primary and secondary outcomes. HR, 95% CI and p-value presented by SBP categories in a crude and adjusted model. ^a^ Adjusted for age, sex, inclusion year, hospital. ^b^ Reference category for Cox proportional hazard regression **Abbreviations** ASCVD: atherosclerotic cardiovascular disease. CI: confidence interval. CHD: coronary heart disease. ICH: intracerebral hemorrhage. HR: hazard ratio. MI: myocardial infarction. SBP: systolic blood pressure

**Figure 4.**
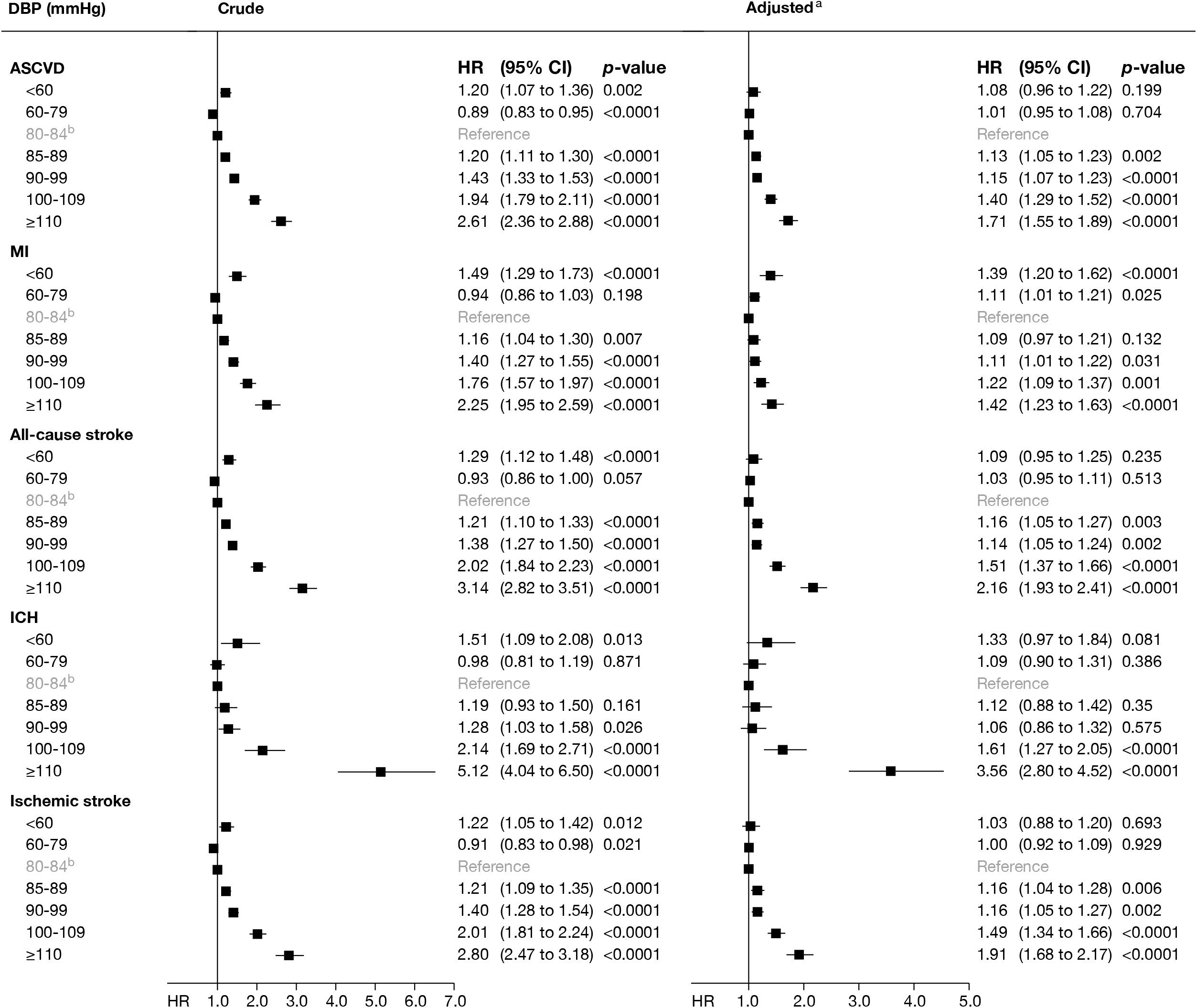
Hazard ratio for primary and secondary outcomes. HR, 95% CI and p-value presented by DBP categories in a crude and adjusted model. ^a^ Adjusted for age, sex, inclusion year, hospital. ^b^ Reference category for Cox proportional hazard regression **Abbreviations** ASCVD: atherosclerotic cardiovascular disease. CI: confidence interval. CHD: coronary heart disease. DBP: diastolic blood pressure ICH: intracerebral hemorrhage. HR: hazard ratio. MI: myocardial infarction.

**Figure 5.**
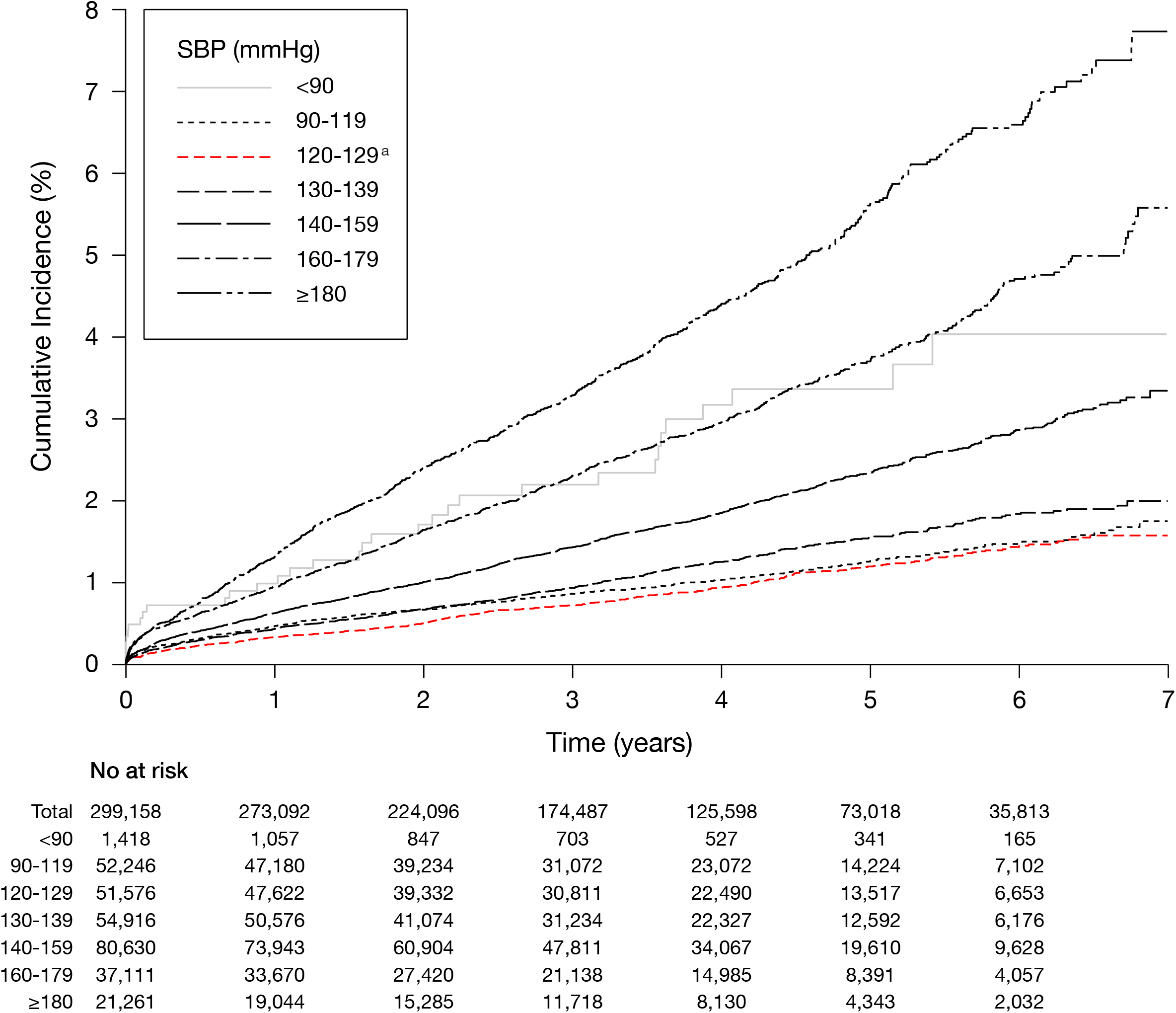
Cumulative incidence of MI. Cumulative incidence are presented by SBP categories. Number at risk presents the number of subjects entering each interval. ^a^ Reference category for Cox proportional hazard regression **Abbreviations** MI: myocardial infarction. SBP: systolic blood pressure

A total of 6,661 (2.2%) all-cause stroke events occurred, and of those were 51.0% male (n=3,470). Patients with DBP ≥110 had more than a twofold association (HR 2.16, 95% CI 1.93-2.41) to a stroke event (Figure 4-5). The six-year cumulative incidence was 1% for the reference category compared to 9% in the highest SBP group (Figure 6).

**Figure 6.**
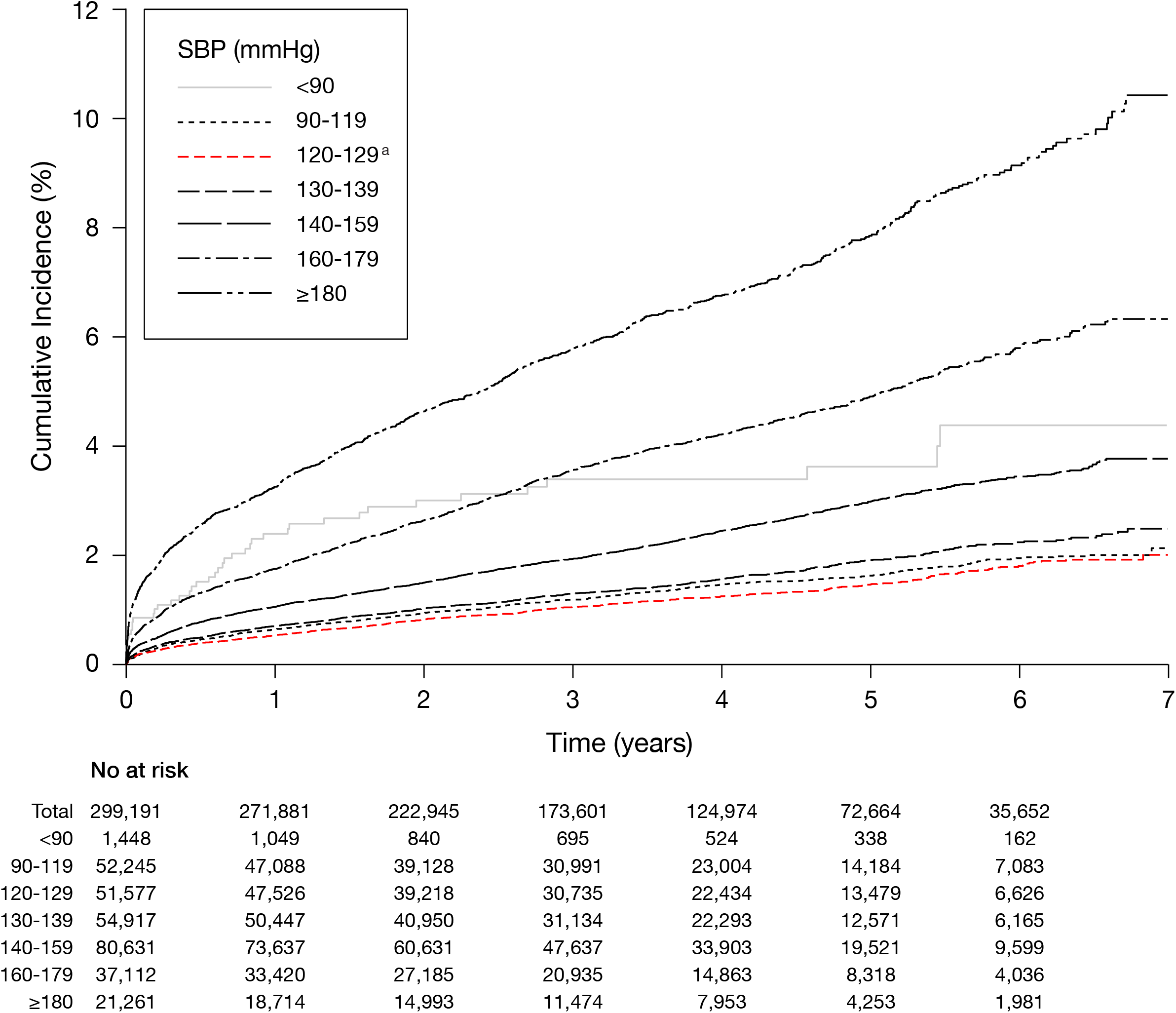
Cumulative incidence of all-cause stroke. Cumulative incidence are presented by SBP categories. Number at risk presents the number of subjects entering each interval. ^a^ Reference category for Cox proportional hazard regression **Abbreviations** SBP: systolic blood pressure

For the endpoints ischemic stroke and intracerebral haemorrhage, a total of 5,355 respectively 1,119 events occurred. The highest HR for ischemic stroke was in the ≥110 mmHg DBP category (HR 1.91, 95% CI 1.68-2.17. The strongest association for intracerebral haemorrhage was in the top DBP category (HR 3.56 95% CI 2.80-4.52) (Figure 4-5).

### Subgroup analyses

Patients with a history of hypertension had 5,667 (7.6%) ASCVD events, compared to 3,300 (1.5%) for those without the condition (Supplementary Table 6). BP above the reference category had a progressively increased association with ASCVD regardless of previous hypertension. Patients with no prior hypertension diagnosis had a lower absolute event rate, but a stronger and steeper relative association with ASCVD compared to the hypertension group. The strongest association for ASCVD was in SBP ≥180 mmHg, for patients with no history of hypertension (HR 2.02, 95% CI 1.75-2.33) (Figure 7).

**Figure 7.**
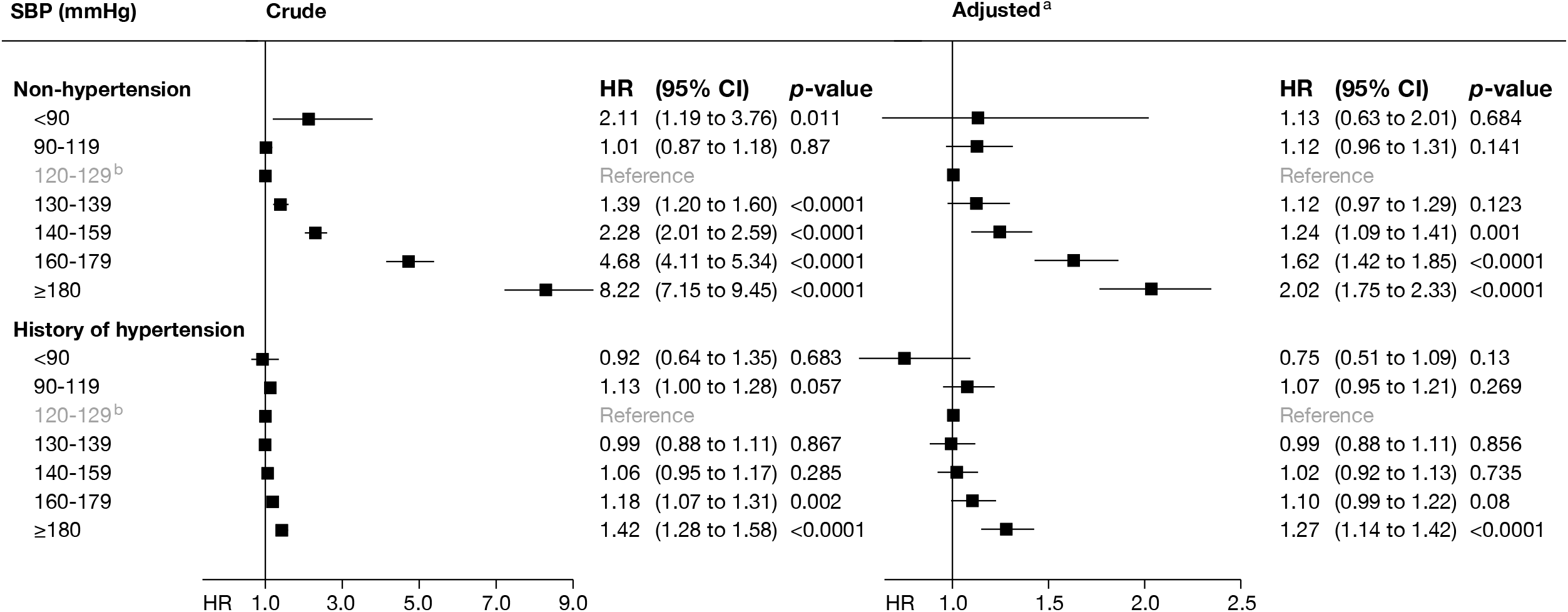
Comparison of hazard ratio for ASCVD by hypertension and non-hypertension groups. HR, 95% CI and p-value presented by SBP categories in a crude and adjusted model. ^a^ Adjusted for age, sex, inclusion year, hospital. ^b^ Reference category for Cox proportional hazard regression **Abbreviations** ASCVD: atherosclerotic cardiovascular disease. CI: confidence interval. HR: hazard ratio.

A total of 76,474 patients (25.5%) were admitted to inpatient care and among these were 50.5% men. The admitted patients had 4,614 ASCVD events during the study period compared to 4,323 events for those who were directly discharged. The mean age for the admitted patients were 60.0±20.4 years and 46.5±18.9 years for the other group. There was a progressively increased hazard ratio among patients with SBP in the categories above the reference group. The association was similar in the two groups in the adjusted model. (Figure 8).

**Figure 8.**
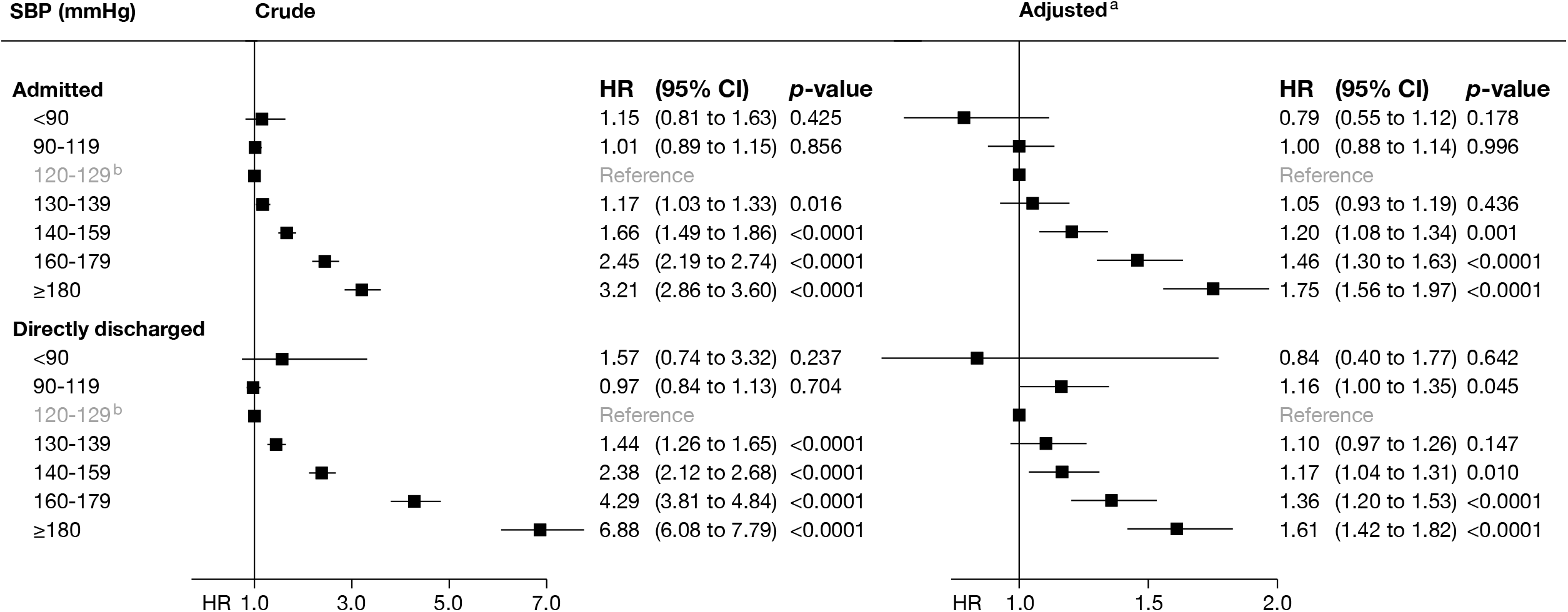
Comparison of hazard ratio for ASCVD by admitted and directly discharged patients. HR, 95% CI and p-value presented by SBP categories in a crude and adjusted model. ^a^ Adjusted for age, sex, inclusion year, hospital. ^b^ Reference category for Cox proportional hazard regression **Abbreviations** ASCVD: atherosclerotic cardiovascular disease. CI: confidence interval. HR: hazard ratio.

The most common chief complaint was in the internal medicine discipline with 3,934 incident ASCVD events (Supplementary Table 7). There was a gradual association above the reference category between BP and incident ASCVD in all chief complaints except for the orthopaedics and otolaryngology disciplines. The strongest association for BP with ASCVD was in the surgery discipline in the crude model (HR 9.35, 95% CI 7.48-11.70). In the adjusted model, the strongest association with ASCVD were in the neurology discipline (Figure 9).

**Figure 9.**
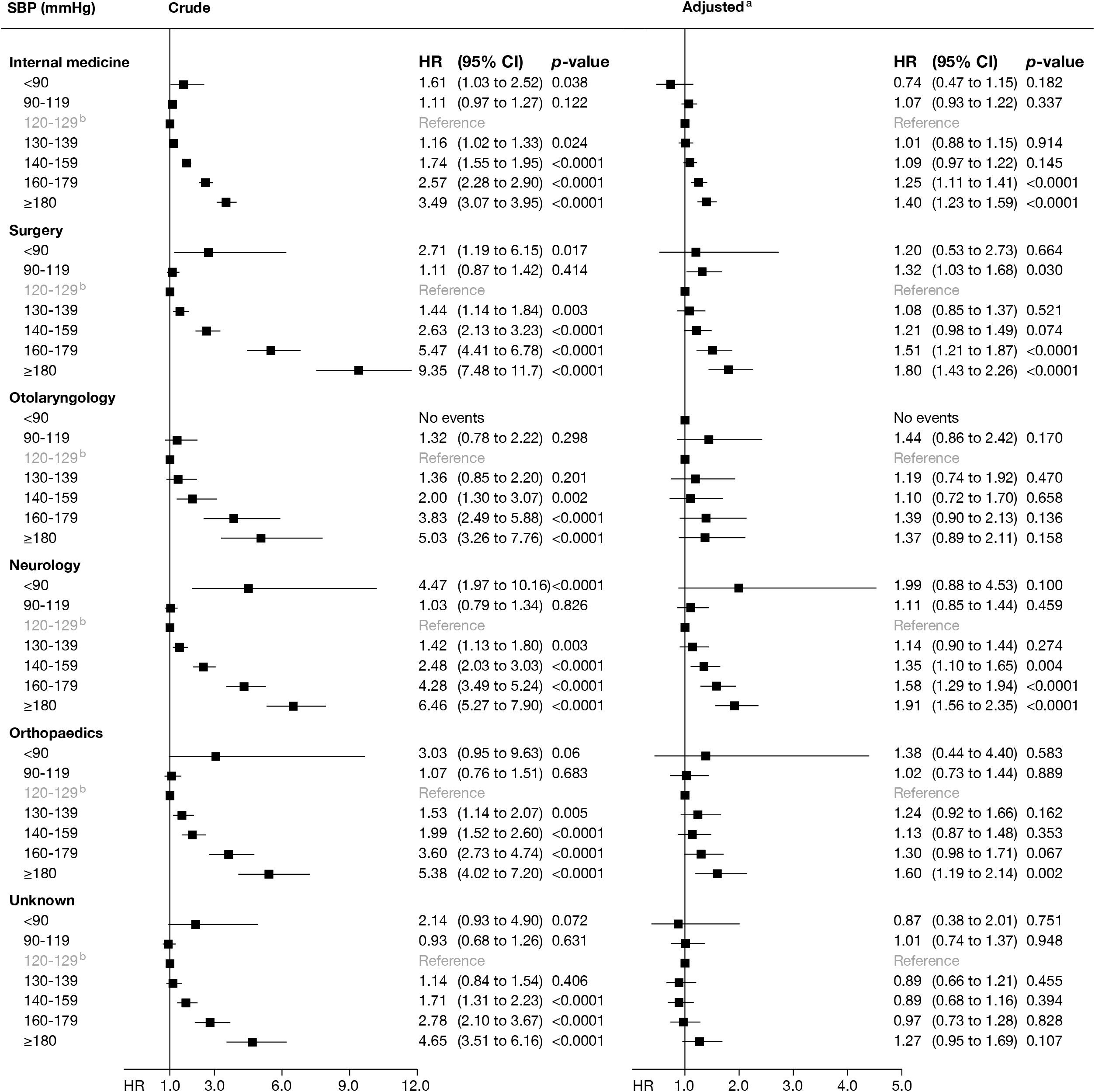
Hazard ratio for ASCVD by chief complaints. HR, 95% CI and p-value presented by SBP categories in a crude and adjusted model. ^a^ Adjusted for age, sex, inclusion year, hospital. ^b^ Reference category for Cox proportional hazard regression **Abbreviations** ASCVD: atherosclerotic cardiovascular disease. CI: confidence interval. HR: hazard ratio. MI: myocardial infarction. SBP: systolic blood pressure

### Number needed to screen

PPE was estimated to 1,966 ASCVD events, 672 CHD, 1,252 strokes. The estimated NNS to find high BP and to prevent one event for ASCVD, CHD and stroke, during a median of 42 months of follow-up, was 151 (95% CI 127-190), 442 (95% CI 360-572), and 157 (95% CI 134-195), respectively, whereas the NNT was estimated to 71, 208, and 112. For patients that were directly discharged, NNS for ASCVD was 216, whereas, NNT for high BP was estimated to 103. To prevent one fatal ASCVD event, 450 patients in the ED needed to be screened for high BP and 150 of those needed to be treated for high BP. (Table 3)

**Table 3.** Number needed to screen and treat to prevent CHD, stroke, and ASCVD. The estimates were based on the assumption that all detected patients with SBP ≥140 mmHg were treated with antihypertensive therapy.

| Estimates for all patients | CHD | Stroke | ASCVD |
| --- | --- | --- | --- |
| Relative risk reduction. % (95% CI) <sup>a</sup> | 22 (17 to 27) | 41 (33 to 48) | 32 (25 to 38) |
| PPE. No (95% CI) | 672 (519 to 824) | 1,252 (1,007 to 1,465) | 1,966 (1,561 to 2,341) |
| PPE per ED patient. % (95% CI) | 0.23 (0.17 to 0.28) | 0.64 (0.51 to 0.75) | 0.66 (0.53 to 0.79) |
| PPE per 1,000 ED patients. No (95% CI) | 3 (2 to 3) | 7 (6 to 8) | 7 (6 to 8) |
| NNT. No (95% CI) | 208 (170 to 269) | 112 (96 to 139) | 71 (60 to 90) |
| NNS. No (95% CI) | 442 (360 to 572) | 157 (134 to 195) | 151 (127 to 190) |
| Estimates for directly discharged | CHD | Stroke | ASCVD |
| Relative risk reduction. % (95% CI) <sup>a</sup> | 22 (17 to 27) | 41 (33 to 48) | 32 (25 to 38) |
| PPE. No (95% CI) | 499 (385 to 612) | 1,225 (986 to 1,435) | 1,366 (1,084 to 1,626) |
| PPE per ED patient. % (95% CI) | 0.17 (0.13 to 0.21) | 0.42 (0.33 to 0.49) | 0.46 (0.37 to 0.55) |
| PPE per 1,000 ED patients. No (95% CI) | 2 (2 to 3) | 5 (4 to 5) | 5 (4 to 6) |
| NNT. No (95% CI) | 280 (228 to 362) | 114 (98 to 142) | 103 (86 to 129) |
| NNS. No (95% CI) | 591 (482 to 765) | 241 (206 to 299) | 216 (182 to 272) |
| Estimates for fatal events | CHD | Stroke | ASCVD |
| Relative risk reduction. % (95% CI) <sup>a</sup> | 22 (17 to 27) | 41 (33 to 48) | 32 (25 to 38) |
| PPE. No (95% CI) | 465 (359 to 571) | 1,148 (924 to 1,344) | 932 (740 to 1,109) |
| PPE per ED patient. % (95% CI) | 0.16 (0.12 to 0.19) | 0.29 (0.23 to 0.34) | 0.22 (0.18 to 0.26) |
| PPE per 1,000 ED patients. No (95% CI) | 2 (2 to 2) | 3 (3 to 4) | 3 (2 to 3) |
| NNT. No (95% CI) | 300 (245 to 388) | 122 (104 to 151) | 150 (126 to 189) |
| NNS. No (95% CI) | 644 (525 to 833) | 346 (295 to 429) | 450 (378 to 567) |

<sup>a</sup>Relative risk reduction based on 10-mmHg SBP reduction, as estimated by Law et al. <sup>8</sup>
**Abbreviations** CI: confidence interval. PPE: potentially preventable event. NNS: number needed to screen. NNT: number needed to treat. SBP: systolic blood pressure.

## DISCUSSION

The main finding in this study was the strong association between BP recorded in the ED and incident ASCVD, irrespective of whether patients were admitted to inpatient care or had a history of hypertension. In patients without a history of hypertension, the association with incident ASCVD was stronger, than for those with an established diagnosis. By using effect sizes for antihypertensive treatment from meta-analysis^8^, we estimated that to prevent one future ASCVD event during a median of 42 months of follow-up, antihypertensive treatment needed to be initiated in 71 patients with an ED-measured SBP ≥140 mmHg. The association with ASCVD was gradually increasing for BP levels that corresponded to hypertension grade 1, 2 and 3^14^ (Supplementary Table 1). Furthermore, a statistically significant association with incident ASCVD was observed for patients with DBP level in the high normal category. Our findings show that hypertension grades may be of interest for the ED setting, and ED-measured BP is of value not only to evaluate the acuity of the patient’s health but also to identify patients at high risk for cardiovascular events in the long-term and therefore provides a tool to prevent them.

Previous population-based research has shown that an increase in SBP results in a substantially increased risk for cardiovascular events, irrespective of baseline BP. ^6,7^ While findings in the present study also showed a strong association between BP and incident ASCVD, the magnitude of the effect was smaller in comparison to the population-based studies. Since the current study did not account for the regression dilution bias due to single BP measurements, our findings probably underestimate the true association between BP in the ED and incident ASCVD.^18^ The influence of different stressors, both related to acute disease and the ED environment, may cause an increase of BP in the ED, and elevated BP levels are commonly observed.^4^ Previously, the prognostic relevance of elevated BP in the ED, and if it can be a basis for decisions to initiate or intensify antihypertensive treatment in this setting has been unknown. However, it should be noted that BP measured during other stress, such as exercise, previously have provided independent prognostic information regarding incident ASCVD.^19,20^ Although it can be speculated that different mechanisms influence BP in the ED and its association with ASCVD, which may explain some of the differences in this study compared to the population-based studies. Here we, for the first time, provide evidence that a first BP obtained in the ED among an unselected population with ED visitors is associated with incident ASCVD.

All secondary endpoints had similar results as for ASCVD. However, there was a remarkably strong association between DBP ≥110 mmHg and incident intracerebral haemorrhage, in the adjusted model, which none of the previously population-based studies has reported. Patients with a history of hypertension had a higher absolute risk to develop ASCVD, but interestingly the association between elevated BP and incident ASCVD was stronger for patients with no history of hypertension. A possible explanation may be that antihypertensive treatment used in the hypertension group attenuated the association.

The estimated NNS was based on the assumption that all detected patients with an SBP ≥140 mmHg were prescribed adequate antihypertensive therapy and succeeded a 10-mmHg SBP-reduction. Prior research has presented a NNS for hypertension with BP-lowering actions,^21^ to the best of our knowledge none have reported the potential benefit of using BPs obtained in the ED with an estimated potential benefit of reduced events. Findings in this study indicate that the ED-setting has great potential to find undiagnosed and undertreated hypertension, which will reduce morbidity and mortality in CVD if effective antihypertensive therapy is initiated.

The main strength of this study was the large cohort size and the long follow-up period, which yielded a strong statistical power. The Swedish National Patient register is well-known for its high-quality and accurate data, with a high reported positive predictive value for most of the diagnoses.^17^ It is noticeable that a few cases had missing data. One potential weakness in this study was the uncertainty of how BP was measured, and due to discrepancies in methods for obtaining BP, deviations for BP may vary from case to case. Preferred end-digits for BP by multiples of 5 and 10 were noticed in the data, which indicates that rounding occurred for some cases. The impact of the rounding was reduced by categorising the BP into groups. Another limitation is that any changes in antihypertensive treatment that were initiated based on elevated ED-measured BP were not considered. Such treatment may have attenuated the association with incident ASCVD and therefore underestimated the true association. We encourage future studies to investigate whether initiation of antihypertensive treatment in the ED yield better long-term prognosis for patients with elevated ED-recorded BP, compared to current praxis.

The clinical value of the findings in this study is that ED-measured BP has a great potential to find undiagnosed hypertension and to prevent future ASCVD events if antihypertensive therapy is initiated. Patients directly discharged from the ED should be assessed more carefully as no further BP recordings take place. One ASCVD event could be prevented for every 107 discharged patients with a high BP if effective antihypertensive therapy is initiated. Thereby, high BP recordings should not be disregarded as isolated events but should result in an initiation of antihypertensive treatment or referral for follow-up evaluation. It is conceivable that the external validity of this study applies to other EDs, particularly considering the broad inclusion criteria, which included all Swedish citizens ≥ 18 years old with a BP obtained in the ED.

In conclusion, findings in this study show that elevated BP, when measured in the ED, is strongly associated with incident ASCVD, MI, and stroke. High BP recordings in EDs should not be disregarded as isolated events, but an opportunity to detect and improve treatment of hypertension. ED-measurements of BP provide an important and under-used tool with great potential to reduce morbidity and mortality associated with hypertension.

## Data Availability

No additional data are available.

## FOOTNOTES

### Contributions

PS conducted the study, acquired all data and the ethical permit from the Regional Ethical Review Board in Stockholm. PO wrote the manuscript, supervised by PS and supported by PHS and HH. HH performed the statistical analysis. HH, PO and PS interpreted the data. PO and HH produced all figures. All authors critically revised the manuscript for intellectual content and approved the final version of the report. The corresponding author had access to all data in this study and is responsible for the decision to submit this study.

### Ethical approval

This study has been approved by the Regional Ethical Review Board in Stockholm, without the requirement of informed consent. Ethical review number 2016/888-31.

### Transparency declaration

The guarantor (PS) affirms that this manuscript is an honest, accurate and transparent account of the study being reported; and that no important aspects of the study have been omitted; and that any discrepancies from the study as planned have been explained.

### Data sharing

No additional data are available

## SUMMARY

### What is already known on this topic

- Hypertension is the leading risk factor for CVD and remains a global challenge in prevention, detection, and improving treatment.
- Antihypertensive treatment reduces the risk of incident ASCVD.
- BP is recorded in most patients in the ED, but the predictive value of BP recorded in the ED for the long-term prognosis is unknown. It is not known whether elevated BP in the ED is associated with ASCVD, MI or stroke.

### What this study adds

- This is the first study to report the association between ED-recorded BP and long-term incident cardiovascular diseases.
- Elevated BP in the ED is progressively associated with incident ASCVD, MI and stroke, for BP levels above normal (SBP: ≥140 mmHg and DBP: ≥90 mmHg)
- The six-year cumulative incidence of ASCVD in SBP ≥180 was 12%, compared to 2% for the normal SBP.
- To prevent one ASCVD event, during a median of 42 months of follow-up, the number needed to screen for high BP in the ED was 151, whereas the number needed to treat with antihypertensive treatment was estimated to 71 patients.
- ED-measurements of BP provide an important and under-used tool with great potential to reduce morbidity and mortality associated with hypertension.

---

**Supplementary Figure 1.**
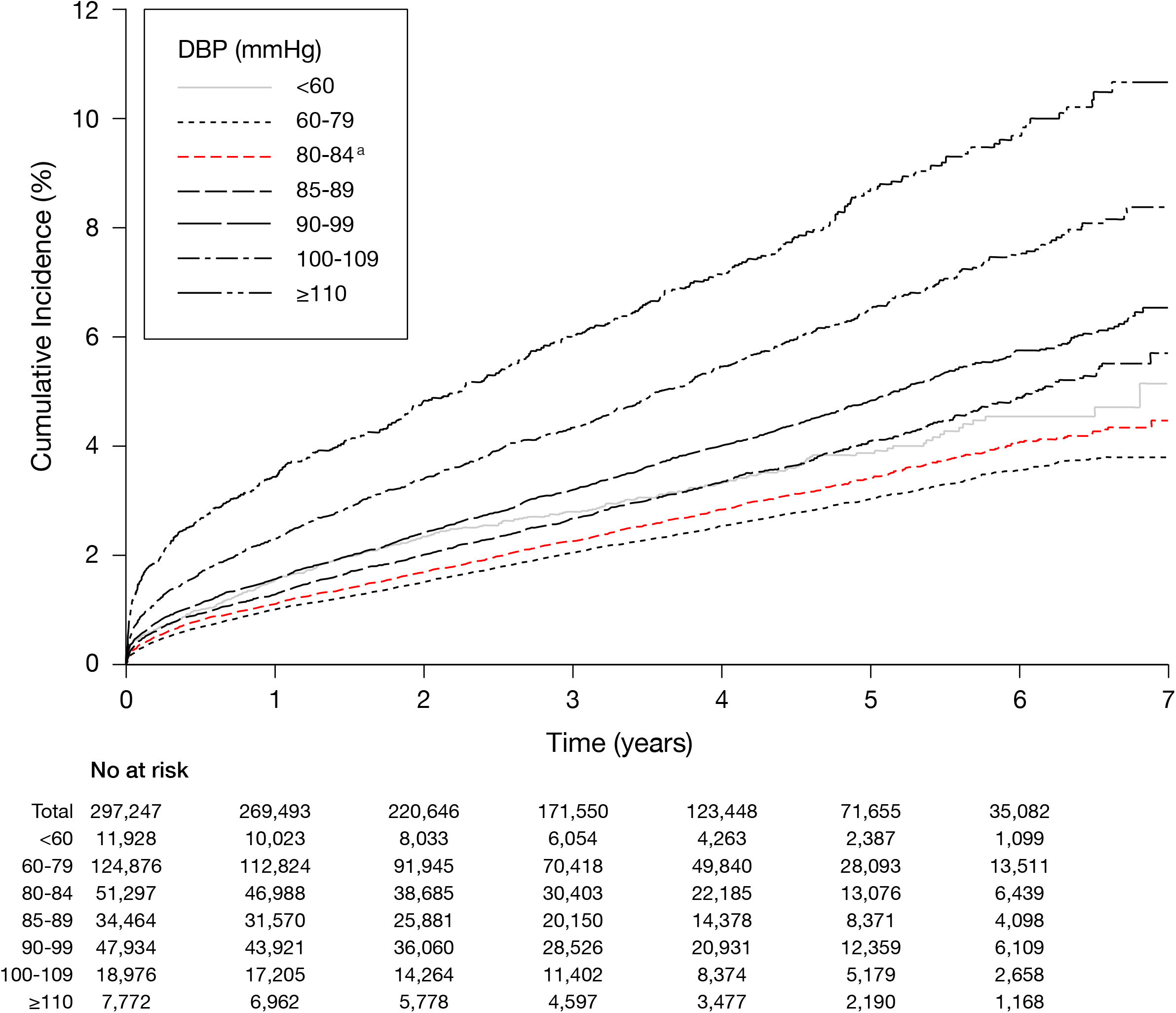
Cumulative incidence of ASCVD. Cumulative incidence are presented by DBP categories. Number at risk presents the number of subjects entering each interval. ^a^ Reference category for Cox proportional hazard regression **Abbreviations** DBP: diastolic blood pressure

**Supplementary Figure 2.**
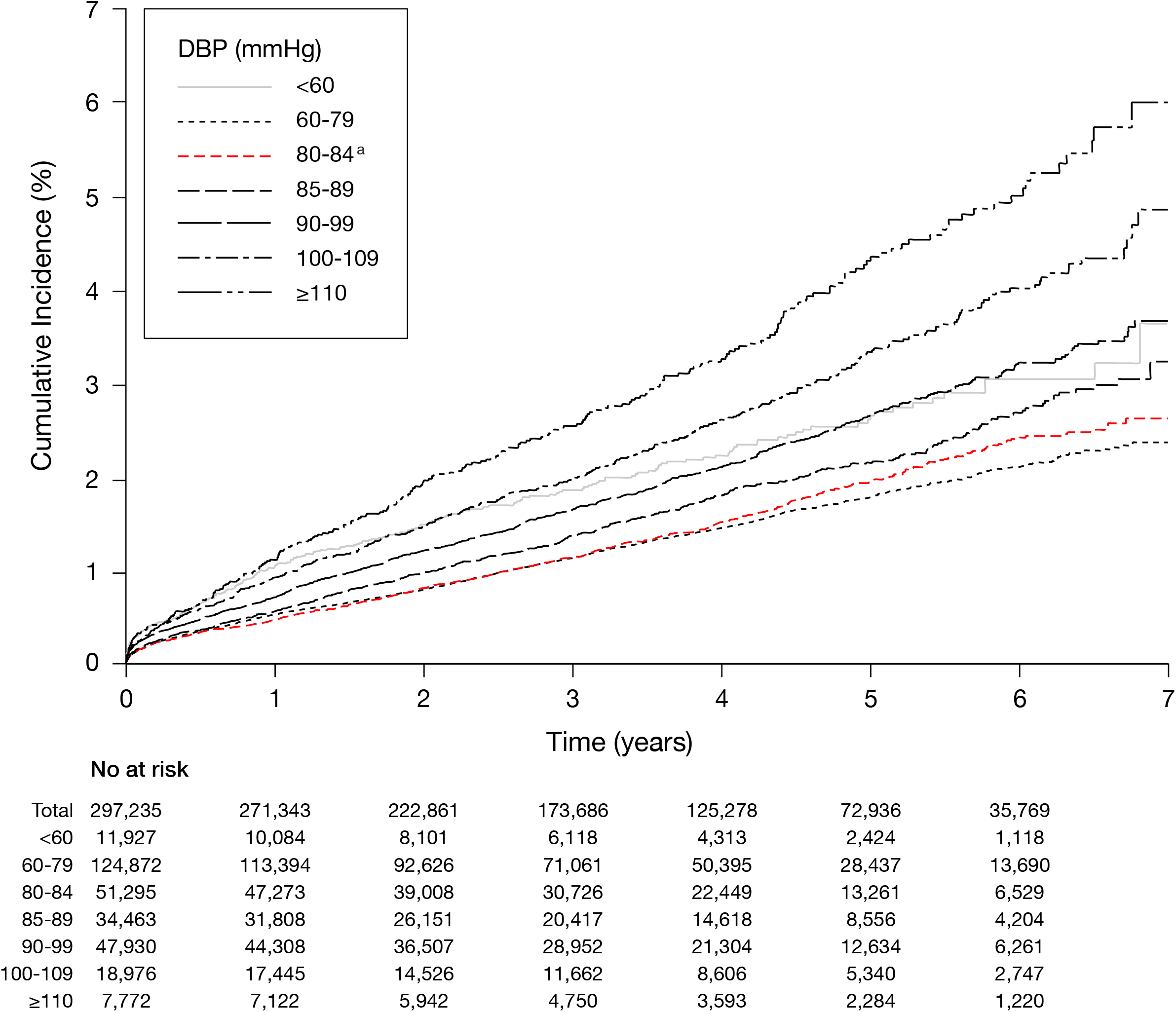
Cumulative incidence of MI. Cumulative incidence are presented by DBP categories. Number at risk presents the number of subjects entering each interval. ^a^ Reference category for Cox proportional hazard regression **Abbreviations** MI: myocardial infarction. DBP: diastolic blood pressure

**Supplementary Figure 3.**
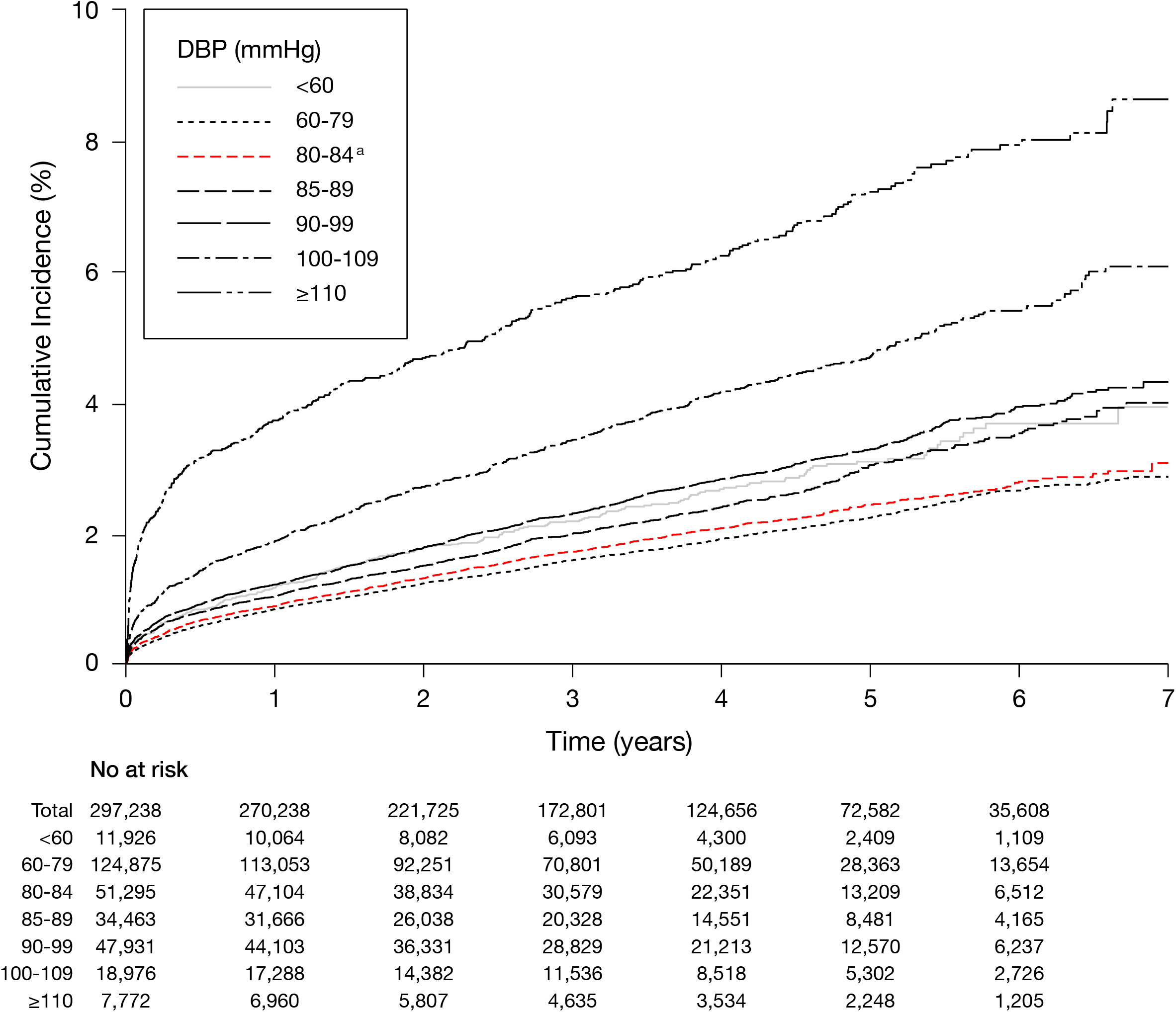
Cumulative incidence of all-cause stroke. Cumulative incidence are presented by DBP categories. Number at risk presents the number of subjects entering each interval.

^a^ Reference category for Cox proportional hazard regression

**Abbreviations** DBP: diastolic blood pressure

